# Disparities in COVID-19 related knowledge, attitudes, beliefs and behaviours by health literacy

**DOI:** 10.1101/2020.06.03.20121814

**Authors:** KJ McCaffery, RH Dodd, E Cvejic, J Ayre, C Batcup, JMJ Isautier, T Copp, C Bonner, K Pickles, B Nickel, T Dakin, S Cornell, MS Wolf

**Author notes:** **Corresponding author:** Professor Kirsten McCaffery School of Public Health. Room 128B Edward Ford Building (A27), The University of Sydney, NSW, 2006.

## Abstract

**Objectives:** To explore the variation in understanding, attitudes and uptake of COVID-19 health advice during the 2020 pandemic lockdown by health literacy.

**Study design:** National cross sectional community survey.

**Setting:** Australian general public.

**Participants:** Adults aged over 18 years (n = 4362).

**Main outcome measures:** Knowledge, attitudes and behaviours related to COVID-19; health literacy and socio-demographic factors.

**Results:** People with inadequate health literacy had poorer understanding of COVID-19 symptoms (49% vs 68%; p<0.001), were less able to identify behaviours to prevent infection (59% vs 72%; p<0.001), and experienced more difficulty finding information and understanding government messaging about COVID-19 than people with adequate health literacy. They were less likely to rate social distancing as important (6.1 vs 6.5, p<0.001) and reported more difficulty remembering/accessing medication since lockdown (3.6 vs 2.7, p<0.001). Importantly there was higher endorsement of misinformation beliefs related to COVID-19 and vaccination in people with lower health literacy. Similar results were observed among people who primarily speak a language other than English at home.

**Conclusion:** Our findings show important disparities by health literacy and language in COVID-19 related knowledge, attitudes and behaviours that have the potential to undermine efforts to reduce viral transmission and may lead to social inequalities in health outcomes in Australia. Those with the greatest burden of chronic disease are most disadvantaged, and most likely to experience severe disease and die from COVID-19. Addressing the health literacy needs of the community in public health messaging about COVID-19 must now be a priority in Australia.

## INTRODUCTION

The coronavirus pandemic represents the biggest public health challenge Australia and the world have faced in living memory. Because COVID-19 spreads so rapidly, the pandemic has placed unprecedented strain on health systems globally. People at greater risk of a severe response to COVID-19 include those aged over 60 years, in aged care facilities, with compromised immune systems (e.g. cancer) and those with chronic medical conditions (1). Although data is still emerging, it suggests that people with chronic disease and multimorbidity are particularly susceptible (2). There are well known social inequalities in chronic disease with higher rates in more disadvantaged populations. Perhaps predictably, evidence of large disparities in COVID-19 deaths by ethnicity and socioeconomic groups has emerged in the US and UK (2, 3).

Currently there are no proven antiviral treatments and no vaccine. This means that to control the spread of COVID-19 we are largely reliant on individual behaviour to comply with restrictions and follow recommended advice on behaviours such as physical distancing, voluntary testing, self-isolation, and hand hygiene. Lockdown measures can enforce some of these behaviours, for example, by restricting travel, requiring returning travellers to self-isolate, closure of public recreational spaces, dramatically limiting individual contacts, and an extensive shift to attend work or school remotely. These combined efforts to slow the spread of COVID-19 have had notable success in Australia. However, different levels of engagement with these lockdown measures within our community may lead to hotspots or even a second wave, and certain groups may be more severely affected by COVID-19.

Since an effective response to the virus requires individuals to modify their behaviour, their engagement with public health information is a pivotal element. This has meant being able to process and understand rapidly evolving public health messages, and then actioning them. It is well known that people vary in their ability to understand, access and action health advice and make informed health decisions –a set of skills commonly called “health literacy”(4). Health literacy has emerged as one of the strongest psychosocial determinants of health outcomes and explains a range of health inequalities by age, ethnicity and socioeconomic position (5). Concern about the quality and appropriateness of communication for people with lower health literacy, and for other vulnerable population subgroups early on in the lockdown campaign, has been expressed (6). This was alongside widespread concern about inconsistent messaging (e.g. on sending children to school) and lack of clarity over key preventive behavioural advice (physical distancing).

We set out to explore the understanding, uptake and impact of COVID-19 health advice during the 2020 pandemic lockdown among a diverse national sample. Our aim was to understand if vulnerable populations might be further disadvantaged in their understanding and attitudes regarding COVID-19 prevention measures. We examined variation in knowledge, attitudes, behaviours and psychosocial outcomes by health literacy and key sociodemographic factors.

## METHODS

### Setting

The survey was carried out in collaboration with a sister survey conducted in the United States in March (7)(Wolf et al 2020). The adapted Australian version of the survey was conducted between April 17^th^-22^nd^ 2020, when lockdown had been in place for 3 weeks.

### Study design

A cross-sectional survey was conducted using the online platform Qualtrics. This study was approved by The University of Sydney Human Research Ethics Committee (2020/212).

### Participants

Participants were aged 18 years and over, able to read and understand English and currently residing in Australia. Participants were recruited via social media (Facebook and Instagram) and Dynata, a large market research company with over 600,000 online panel members. Participants recruited via social media were given the opportunity to enter into a prize draw for the chance to win one of ten $20 gift cards upon completion of the survey. Participants recruited via Dynata received points for completing the survey, which can be redeemed for gift vouchers, donations to charities or money.

### Measurements

Sociodemographic variables including age, gender, educational status, employment status, country of birth, area of residence, number in household, primary language spoken at home, and self-identification as Aboriginal or Torres Strait Islander were collected. In addition, health insurance status, self-reported chronic diseases and self-reported overall health were obtained. Changes in consumption of unhealthy snacks and alcohol intake was assessed (8). We assessed health literacy using Health Literacy single item screener (8) and eHealth Literacy Scale (eHeals) (9), and numeracy using the Subjective Numeracy Scale (10). The Consumer Health Activation Index (CHAI) was used to determine patient activation (11). Anxiety and depression were measured using self-reported history of anxiety, depression and the State-Trait Anxiety Inventory (STAI) (12). Participants were asked to indicate their awareness and concerns (7), perceived financial impact (13), knowledge, sources of information, personal preparedness (14), behaviour change, daily impact and support for misinformation of COVID-19 (15) (Box 1 in the Appendix lists the items).

### Statistical analysis

Descriptive statistics were calculated for all participant characteristics (Table 1) and study outcome measures (Supplementary Table S1). Associations between key participant characteristics (see Tables 1–2) and outcomes were examined in univariable analyses using X^2^ tests, t-tests, or analysis of variance (as appropriate). To explore variation in outcomes by health literacy adequacy, multivariable linear regression models were used to estimate marginal means (with 95% confidence intervals) for continuous outcomes, and generalised linear models with a modified Poisson approach (16) to estimate relative risks (with 95% confidence intervals) for dichotomous outcomes. All multivariable models controlled for age group, gender, number of chronic health conditions, language spoken at home, private health insurance status, and employment status. Statistical analyses were conducted in Stata/IC v16.1 (StataCorp, College Station, Texas, USA).

**Table 1.**
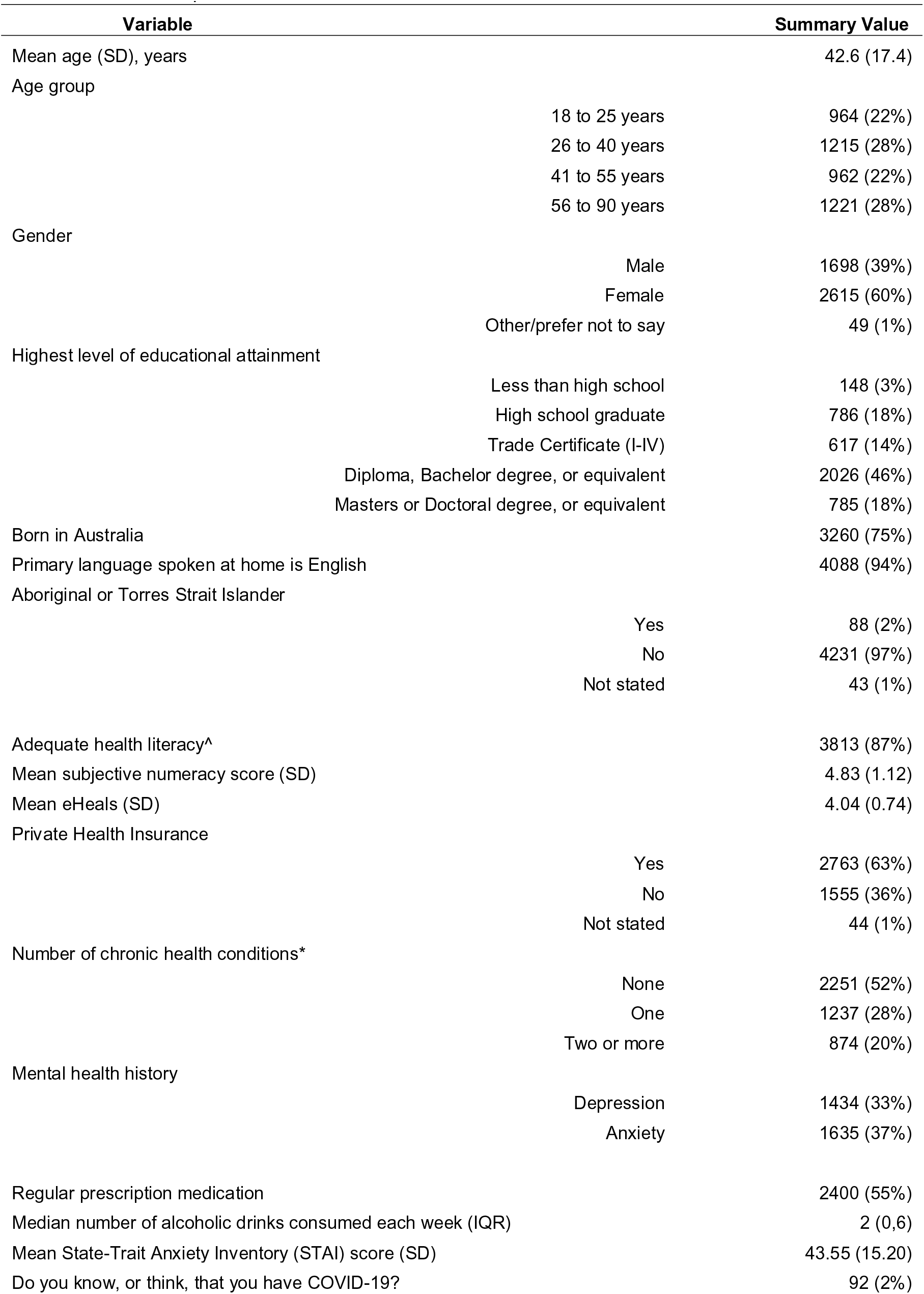
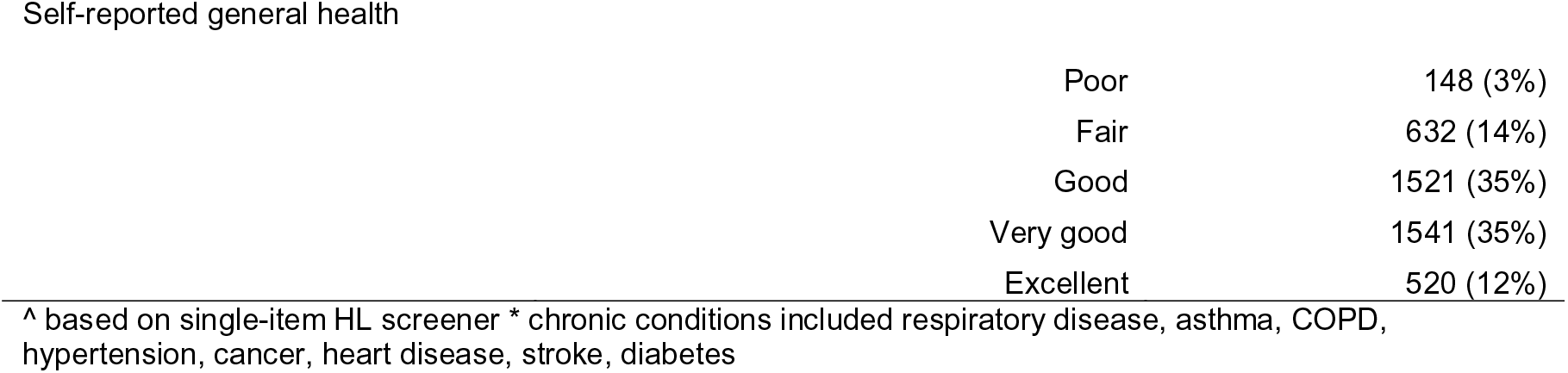
Descriptive characteristics of analysis sample (N=4362). Data are shown as n (%) unless otherwise specified.

| Variable | Summary Value |
| --- | --- |
| Mean age (SD), years | 42.6 (17.4) |
| Age group |  |
| 18 to 25 years | 964 (22%) |
| 26 to 40 years | 1215 (28%) |
| 41 to 55 years | 962 (22%) |
| 56 to 90 years | 1221 (28%) |
| Gender |  |
| Male | 1698 (39%) |
| Female | 2615 (60%) |
| Other/prefer not to say | 49 (1%) |
| Highest level of educational attainment |  |
| Less than high school | 148 (3%) |
| High school graduate | 786 (18%) |
| Trade Certificate (I-IV) | 617 (14%) |
| Diploma, Bachelor degree, or equivalent | 2026 (46%) |
| Masters or Doctoral degree, or equivalent | 785 (18%) |
| Born in Australia | 3260 (75%) |
| Primary language spoken at home is English | 4088 (94%) |
| Aboriginal or Torres Strait Islander |  |
| Yes | 88 (2%) |
| No | 4231 (97%) |
| Not stated | 43 (1%) |
| Adequate health literacy^ | 3813 (87%) |
| Mean subjective numeracy score (SD) | 4.83 (1.12) |
| Mean eHeals (SD) | 4.04 (0.74) |
| Private Health Insurance |  |
| Yes | 2763 (63%) |
| No | 1555 (36%) |
| Not stated | 44 (1%) |
| Number of chronic health conditions* |  |
| None | 2251 (52%) |
| One | 1237 (28%) |
| Two or more | 874 (20%) |
| Mental health history |  |
| Depression | 1434 (33%) |
| Anxiety | 1635 (37%) |
| Regular prescription medication | 2400 (55%) |
| Median number of alcoholic drinks consumed each week (IQR) | 2 (0,6) |
| Mean State-Trait Anxiety Inventory (STAI) score (SD) | 43.55 (15.20) |
| Do you know, or think, that you have COVID-19? | 92 (2%) |
| Self-reported general health |  |
| Poor | 148 (3%) |
| Fair | 632 (14%) |
| Good | 1521 (35%) |
| Very good | 1541 (35%) |
| Excellent | 520 (12%) |

<sup>^</sup> based on single-item HL screener \* chronic conditions included respiratory disease, asthma, COPD, hypertension, cancer, heart disease, stroke, diabetes

**Table 2.** Knowledge, attitudes, beliefs and behaviours related to COVID-19 across sample demographics.

| Variable |  | Awareness and concern |  |  |  |  | Knowledge and information |  |  |  |
| --- | --- | --- | --- | --- | --- | --- | --- | --- | --- | --- |
|  |  | Mean seriousness of threat (SD) 1-10 | Not nervous/ Stressed (%) | Not likely to get sick (%) | Mean STAI (SD) | Mean Financial Toxicity 1-5 | Symptoms (%) | Prevention (%) | Mean difficulty finding information 1-10 | Mean difficulty understanding government messaging 1-10 |
| **Age group** |  |  |  |  |  |  |  |  |  |  |
| | 18 to 25 | 7.3 (2.2)\$ | 15.8\$ | 23.1\$ | 48.4 (14.6)\$ | 3.3 (1.1)\$ | 58.0\$ | 66.4\$ | 4.4 (2.4)\$ | 4.8 (2.7)\$ |
|  | 26 to 40 | 7.5 (2.2) | 11.9 | 22.4 | 46.9 (14.6) | 3.1 (1.0) | 62.8 | 72.2 | 4.2 (2.5) | 4.9 (2.8) |
|  | 41 to 55 | 7.8 (2.2) | 16.4 | 21.2 | 43.0 (14.8) | 2.9 (1.0) | 64.2 | 76.5 | 3.7 (2.5) | 4.4 (2.8) |
|  | 56 to 90 | 8.0 (2.1) | 30.1 | 33.7 | 36.7 (14.1) | 2.7 (0.9) | 56.4 | 67.9 | 3.2 (2.3) | 3.6 (2.6) |
| **Gender** |  |  |  |  |  |  |  |  |  |  |
| | Male | 7.3 (2.3)\$ | 27.0\$ | 31.9\$ | 41.3 (15.0)\$ | 3.0 (1.0) | 53.1\$ | 60.9\$ | 3.8 (2.5) | 4.2 (2.8)# |
|  | Female | 7.9 (2.0) | 13.7 | 21.5 | 44.9 (15.1) | 3.0 (1.0) | 64.9 | 76.9 | 3.9 (2.4) | 4.5 (2.8) |
| **Health literacy** |  |  |  |  |  |  |  |  |  |  |
| | Inadequate | 7.1 (2.3)\$ | 16.4 | 30.8# | 48.8 (14.3)\$ | 3.3 (0.9)\$ | 49.4\$ | 58.8\$ | 4.6 (2.4)\$ | 5.0 (2.7)\$ |
|  | Adequate | 7.7 (2.1) | 19.2 | 24.7 | 42.8 (15.2) | 2.9 (1.0) | 61.8 | 72.4 | 3.7 (2.4) | 4.3 (2.8) |
| **Education** |  |  |  |  |  |  |  |  |  |  |
| | High school or less | 7.4 (2.3)\$ | 23.5\$ | 33.1\$ | 44.4 (15.9)# | 3.1 (1.0)\$ | 52.6\$ | 66.3\$ | 4.0 (2.4)^ | 4.3 (2.8) |
|  | Certificate I-IV | 7.5 (2.3) | 21.7 | 28.7 | 44.8 (15.5) | 3.1 (1.1) | 60.1 | 68.1 | 4.1 (2.5) | 4.5 (2.9) |
|  | University | 7.8 (2.1) | 16.7 | 22.2 | 43.0 (14.9) | 2.9 (1.0) | 62.9 | 72.7 | 3.8 (2.4) | 4.4 (2.8) |
| **General health** |  |  |  |  |  |  |  |  |  |  |
| | Poor to Fair | 7.8 (2.3)^ | 15.5# | 22.7^ | 49.4 (16.0)\$ | 3.3 (1.0)\$ | 57.3 | 69.6 | 4.0 (2.5)^ | 4.8 (2.9)\$ |
|  | Good to Excellent | 7.6 (2.2) | 19.6 | 26.1 | 42.3 (14.7) | 2.9 (1.0) | 60.9 | 70.9 | 3.8 (2.4) | 4.3 (2.8) |
| **Chronic health conditions** |  |  |  |  |  |  |  |  |  |  |
| | None | 7.5 (2.2)\$ | 18.1 | 26.2 | 44.2 (14.9)^ | 3.0 (1.0)^ | 61.4 | 71.1 | 3.9 (2.4) | 4.4 (2.7) |
|  | One | 7.7 (2.1) | 20.2 | 25.3 | 42.8 (15.0) | 2.9 (1.0) | 59.2 | 71.1 | 3.8 (2.4) | 4.4 (2.9) |
|  | Two or more | 8.1 (2.1) | 18.9 | 23.9 | 43.0 (16.0) | 3.0 (1.0) | 58.8 | 69.2 | 3.9 (2.6) | 4.3 (2.9) |
| **Language at home** |  |  |  |  |  |  |  |  |  |  |
| | English | 7.7 (2.2)^ | 19.3# | 24.8\$ | 43.4 (15.2)^ | 3.0 (1.0)\$ | 60.4 | 72.0\$ | 3.8 (2.5) | 4.5 (2.8)\$ |
|  | Other | 7.4 (2.3) | 12.8 | 35.8 | 45.5 (14.5) | 3.4 (1.1) | 58.4 | 50.7 | 4.1 (2.5) | 3.7 (2.6) |
| **Private Health Insurance** |  |  |  |  |  |  |  |  |  |  |
| | Yes | 7.7 (2.1)# | 17.6# | 23.5\$ | 43.0 (15.0)# | 2.9 (1.0)\$ | 62.3\$ | 72.0\$ | 3.8 (2.5) | 4.3 (2.8)\$ |
|  | No / not sure | 7.5 (2.3) | 21.0 | 29.0 | 44.5 (15.5) | 3.1 (1.1) | 56.7 | 68.4 | 3.9 (2.5) | 4.6 (2.9) |
| **Employment** |  |  |  |  |  |  |  |  |  |  |
| | Paid employment | 7.6 (2.1) | 15.9\$ | 21.7\$ | 43.4 (14.6) | 2.9 (1.0)\$ | 65.1\$ | 74.6\$ | 3.9 (2.5)^ | 4.5 (2.8)# |
|  | Other | 7.7 (2.2) | 22.0 | 29.6 | 43.8 (15.8) | 3.1 (1.1) | 55.0 | 66.4 | 3.8 (2.4) | 4.3 (2.8) |
| **Change in employment** |  |  |  |  |  |  |  |  |  |  |
| | No change | 7.7 (2.2) | 21.8\$ | 26.1 | 41.6 (14.8)\$ | 2.6 (0.9)\$ | 60.5 | 72.1# | 3.7 (2.4)\$ | 4.2 (2.7)\$ |
|  | Any change | 7.6 (2.2) | 13.6 | 24.4 | 47.0 (15.2) | 3.6 (1.0) | 59.8 | 68.0 | 4.2 (2.5) | 4.8 (2.9) |
| **Living arrangement** |  |  |  |  |  |  |  |  |  |  |
| | Alone | 7.5 (2.4)^ | 25.0\$ | 26.7 | 42.9 (16.0)^ | 2.8 (1.0)\$ | 57.8# | 71.6\$ | 3.6 (2.5)# | 4.2 (2.8) |
|  | Others (with children) | 7.8 (2.1) | 16.5 | 23.2 | 42.7 (14.4) | 3.0 (1.0) | 64.3 | 76.0 | 3.8 (2.4) | 4.5 (2.8) |
|  | Other (No children) | 7.6 (2.2) | 18.6 | 26.2 | 44.1 (15.3) | 3.0 (1.1) | 59.0 | 68.1 | 3.9 (2.5) | 4.4 (2.8) |

^P<0.05; #P<0.01; \$P<0.001

**Table 2 (cont.).** Knowledge, attitudes, beliefs and behaviours related to COVID-19 across sample demographics.
| Variable | Preparedness and response |  |  | Behaviours and impact |  |  |  |  |  | COVID-19/ Vaccination Misinformation Beliefs |  |  |  |
| --- | --- | --- | --- | --- | --- | --- | --- | --- | --- | --- | --- | --- | --- |
|  | Confidence in federal government (%) | Confidence in local / state government (%) | Not personally prepared (%) | Changed daily routine (%) | Changed plans (%) | Mean social distancing score 1-7 | Mean difficulty remembering / accessing medication 1-10 | More alcohol | More snacks | Vaccine effectiveness made-up (%) | COVID-19 threat greatly exaggerated (%) | Herd immunity beneficial and is covered up (%) | Government restrictions stronger than needed (%) |
| **Age group** |  |  |  |  |  |  |  |  |  |  |  |  |  |
| 18 to 25 | 65.8 | 74.6 | 11.1# | 88.4\$ | 95.0\$ | 6.4 (0.8)\$ | 3.9 (2.9)\$ | 25.7\$ | 47.6\$ | 19.4 | 21.2\$ | 22.4\$ | 13.2\$ |
| 26 to 40 | 63.0 | 74.2 | 10.6 | 87.1 | 93.7 | 6.4 (0.8) | 3.3 (2.8) | 33.1 | 46.0 | 18.0 | 17.1 | 16.0 | 17.0 |
| 41 to 55 | 64.8 | 76.5 | 8.0 | 84.3 | 91.9 | 6.5 (0.7) | 2.9 (2.5) | 30.0 | 35.4 | 16.0 | 11.0 | 12.8 | 12.9 |
| 56 to 90 | 67.3 | 76.3 | 7.0 | 80.0 | 89.5 | 6.5 (0.7) | 2.1 (2.0) | 16.1 | 21.3 | 15.8 | 7.0 | 9.7 | 11.3 |
| **Gender** |  |  |  |  |  |  |  |  |  |  |  |  |  |
| Male | 67.0 | 76.1 | 9.2 | 83.8 | 90.6\$ | 6.3 (0.8)\$ | 2.7 (2.5) | 22.7\$ | 31.6\$ | 19.7# | 19.1\$ | 18.0\$ | 18.0\$ |
| Female | 64.6 | 75.2 | 9.1 | 85.5 | 93.6 | 6.5 (0.7) | 2.9 (2.5) | 28.1 | 40.7 | 15.8 | 10.4 | 13.1 | 10.9 |
| **Health literacy** |  |  |  |  |  |  |  |  |  |  |  |  |  |
| Inadequate | 66.3 | 72.9 | 14.6\$ | 77.6\$ | 89.3# | 6.1 (1.0)\$ | 3.6 (2.8)\$ | 20.4# | 37.9 | 26.0\$ | 21.5\$ | 21.1\$ | 19.1\$ |
| Adequate | 65.1 | 75.7 | 8.4 | 85.8 | 92.9 | 6.5 (0.7) | 2.7 (2.5) | 26.8 | 37.0 | 16.0 | 12.7 | 14.1 | 12.9 |
| **Education** |  |  |  |  |  |  |  |  |  |  |  |  |  |
| High school or less | 69.3^ | 74.9 | 9.7# | 81.5\$ | 87.7\$ | 6.4 (0.8)# | 2.8 (2.5) | 21.2\$ | 36.3 | 20.9\$ | 14.2 | 16.4 | 11.3^ |
| Certificate I-IV | 64.8 | 73.6 | 12.2 | 79.7 | 88.2 | 6.4 (0.7) | 3.0 (2.7) | 21.7 | 37.1 | 19.3 | 15.2 | 16.7 | 13.1 |
| University | 64.0 | 75.9 | 8.3 | 87 | 94.9 | 6.5 (0.7) | 2.8 (2.5) | 28.6 | 37.4 | 15.6 | 13.4 | 14.1 | 14.5 |
| **General health** |  |  |  |  |  |  |  |  |  |  |  |  |  |
| Poor to Fair | 52.6\$ | 66.3\$ | 11.0^ | 80.4\$ | 88.1\$ | 6.4 (0.8) | 3.4 (2.8)\$ | 19.1\$ | 38.2 | 18.1 | 10.6# | 11.0\$ | 12.2 |
| Good to Excellent | 68.0 | 77.4 | 8.7 | 85.7 | 93.4 | 6.5 (0.7) | 2.6 (2.4) | 27.5 | 36.9 | 17.1 | 14.5 | 15.8 | 14.0 |
| **Chronic health conditions** |  |  |  |  |  |  |  |  |  |  |  |  |  |
| None | 66.2^ | 76.0^ | 8.9 | 84.1 | 92.4 | 6.4 (0.8)\$ | 2.9 (2.5) | 28.1\$ | 38.8^ | 17.3 | 16.3\$ | 16.6# | 15.3# |
| One | 66.5 | 76.7 | 9.2 | 86.2 | 92.2 | 6.5 (0.7) | 2.7 (2.5) | 26.1 | 36.7 | 17.3 | 11.9 | 14.4 | 11.6 |
| Two or more | 60.9 | 71.9 | 9.7 | 84.6 | 92.9 | 6.5 (0.8) | 2.8 (2.6) | 20.5 | 33.2 | 17.2 | 10.3 | 11.6 | 12.2 |
| **Language at home** |  |  |  |  |  |  |  |  |  |  |  |  |  |
| English | 64.8^ | 75.4 | 9.2 | 84.5 | 92.4 | 6.5 (0.7)\$ | 2.8 (2.5) | 26.7\$ | 37.0 | 16.3\$ | 13.1\$ | 14.6^ | 13.3# |
| Other | 72.3 | 75.5 | 8.4 | 88.3 | 92.3 | 6.3 (0.9) | 3.3 (2.7) | 15.7 | 39.2 | 32.1 | 24.8 | 19.7 | 19.3 |
| **Private Health Insurance** |  |  |  |  |  |  |  |  |  |  |  |  |  |
| Yes | 66.4^ | 77.2\$ | 8.0\$ | 87.0\$ | 94.8\$ | 6.5 (0.7)^ | 2.7 (2.5) | 27.6# | 36.4 | 16.0# | 13.2 | 15.3 | 13.1 |
| No / not sure | 63.1 | 72.2 | 11.1 | 80.9 | 88.4 | 6.4 (0.8) | 2.9 (2.6) | 23.3 | 38.3 | 19.5 | 14.9 | 14.3 | 14.5 |
| **Employment** |  |  |  |  |  |  |  |  |  |  |  |  |  |
| Paid employment | 66.4 | 77.6\$ | 8.3^ | 87.1\$ | 94.7\$ | 6.4 (0.7) | 2.9 (2.6) | 30.7\$ | 38.6^ | 15.8^ | 14.4 | 15.1 | 14.3 |
| Other | 63.9 | 73.0 | 10.0 | 82.2 | 90.0 | 6.5 (0.8) | 2.8 (2.5) | 21.0 | 35.5 | 18.8 | 13.2 | 14.8 | 12.9 |
| **Change in employment** |  |  |  |  |  |  |  |  |  |  |  |  |  |
| No change | 66.6^ | 76.5^ | 7.7\$ | 81.8\$ | 90.9\$ | 6.5 (0.7)^ | 2.6 (2.4)\$ | 23.8\$ | 34.5\$ | 15.5\$ | 11.9\$ | 12.6\$ | 12.1\$ |
| Any change | 62.8 | 73.4 | 11.8 | 90.1 | 95.1 | 6.4 (0.8) | 3.4 (2.8) | 29.9 | 41.7 | 20.3 | 17.3 | 19.0 | 16.3 |
| **Living arrangement** |  |  |  |  |  |  |  |  |  |  |  |  |  |
| Alone | 63.5 | 74.3 | 9.2 | 80.3\$ | 89.4^ | 6.4 (0.7) | 2.7 (2.5) | 23.4# | 32.5^ | 13.3^ | 12.9 | 11.7 | 13.3 |
| Others (with children) | 66.6 | 76.4 | 8.6 | 87.4 | 92.7 | 6.5 (0.7) | 2.8 (2.5) | 29.6 | 37.0 | 17.9 | 13.5 | 15.3 | 14.1 |
| Other (No children) | 65.0 | 75.1 | 9.4 | 84.6 | 93.0 | 6.5 (0.7) | 2.9 (2.6) | 25.0 | 38.2 | 17.8 | 14.2 | 15.5 | 13.5 |

^P<0.05; #P<0.01; \$P<0.001

Sample size was calculated to achieve a specified level of precision in estimates at a 6-month assessment in the prospective cohort, accounting for potential loss to follow-up with each assessment wave.

## RESULTS

We had a total of 4,362 respondents. Sample characteristics are summarised in Table 1. The mean age was 42.6 years (SD 17.4; range: 18–90 years) with 60% female respondents. Most participants (75%) were born in Australia, with 94% speaking English as their primary language at home, 35% had no tertiary qualifications and 36% did not have private health insurance. The presence of at least one chronic health condition was reported by 48% of the sample. Inadequate health literacy (assessed by the SILS) was reported by 13% of the sample.

### COVID-19 Awareness and Concern

Awareness and concern about COVID-19 across sample demographics are shown in Table 2. Notably, older participants (aged 56 to 90 years) rated the seriousness of the threat of COVID-19 as higher than younger participants, but also reported being less nervous, had lower anxiety, and a greater proportion believed they were not likely to get sick than in younger age groups. The perceived seriousness of the threat also increased with number of chronic health conditions reported. Participants who reported speaking a language other than English (LOTE) at home rated the threat of COVID-19 lower, with a greater proportion indicating that they were not likely to get sick compared to those who primarily spoke English at home.

Multivariable analyses examining differences in outcomes by health literacy after adjusting for other sociodemographic variables are displayed in Table 3 (full model estimates are provided in Supplementary Tables S2-S5). Compared to participants with adequate health literacy, individuals with inadequate health literacy rated the seriousness of the threat to be significantly lower (p<0.001), had higher anxiety (p< 0.001), and reported COVID-19 to have a greater impact on their financial situation (p<.001). Participants with inadequate health literacy were more likely (p = 0.018) to think that they would not get sick from COVID-19 compared to participants with adequate health literacy.

**Table 3.** Multivariable^^^ regression models exploring variation in outcomes by health literacy adequacy. Estimates are presented as adjusted risk ratios or marginal mean differences (95% confidence intervals) for individuals with inadequate health literacy relative to individuals with adequate health literacy.

| Outcome | Adjusted risk ratio<br>(95% confidence interval) | Test statistics |
| --- | --- | --- |
| **Awareness and concern** |  |  |
| Perceived seriousness of threat<sup>&</sup> | -0.42 (-0.61 to -0.22) | t(4248)=4.09, $p<0.001$ |
| Not nervous/ stressed | 0.85 (0.70 to 1.03) | $\chi^2(1)=2.47$, $p=0.12$ |
| Not likely to get sick | 1.19 (1.03 to 1.37) | $\chi^2(1)=5.62$, $p=0.018$ |
| Anxiety (STAI)<sup>&</sup> | 4.19 (2.85 to 5.52) | t(4256)=6.14, $p<0.001$ |
| Financial impact<sup>&</sup> | 0.26 (0.13 to 0.32) | t(4302)=4.85, $p<0.001$ |
| **Knowledge and information** |  |  |
| Knowledge of 3 key symptoms | 0.82 (0.75 to 0.90) | $\chi^2(1)=16.92$, $p<0.001$ |
| Knowledge of 3 prevention methods | 0.86 (0.80 to 0.93) | $\chi^2(1)=14.56$, $p<0.001$ |
| Difficulty understanding government messaging<sup>&</sup> | 0.61 (0.35 to 0.86) | t(4302)=4.66, $p<0.001$ |
| **Behaviours and impact** |  |  |
| Not personally prepared | 1.56 (1.22 to 1.99) | $\chi^2(1)=12.51$, $p<0.001$ |
| Changed plans | 0.96 (0.93 to 0.99) | $\chi^2(1)=6.07$, $p=0.014$ |
| Social distancing score<sup>&</sup> | -0.35 (-0.42 to -0.28) | t(4170)=9.75, $p<0.001$ |
| Difficulty remembering/ accessing medication<sup>&*</sup> | 0.44 (0.10 to 0.79) | t(2353)=2.50, $p=0.013$ |
| More alcohol | 0.80 (0.67 to 0.96) | $\chi^2(1)=6.01$, $p=0.014$ |
| More snacking | 0.93 (0.82 to 1.04) | $\chi^2(1)=1.61$, $p=0.20$ |
| **COVID-19/ Vaccination Misinformation Beliefs** |  |  |
| Vaccine effectiveness made-up | 1.45 (1.22 to 1.72) | $\chi^2(1)=17.75$, $p<0.001$ |
| COVID-19 threat greatly exaggerated | 1.24 (1.02 to 1.50) | $\chi^2(1)=4.75$, $p=0.029$ |
| Herd immunity for COVID-19 is beneficial and this fact is being covered up | 1.25 (1.03 to 1.51) | $\chi^2(1)=5.29$, $p=0.022$ |
| Government restrictions are stronger than needed | 1.35 (1.11 to 1.65) | $\chi^2(1)=8.70$, $p=0.003$ |

<sup>^</sup> all models adjusted for age group, gender, chronic health conditions, language spoken at home, private health insurance, and employment status. Full models are provided in Supplementary Table S2-S5; <sup>&</sup> estimates provided are marginal mean differences; \* includes only $n=2364$ who reported taking regular prescription medication.

### COVID-19 knowledge and information

On average, participants estimated that 61.3% of people who get COVID-19 will have only mild symptoms, and that approximately 6.5% of people who are infected with COVID-19 in Australia will die as a result (Supplementary Table S1). Most participants (60%) were able to provide at least three key symptoms of COVID-19, and more than two-thirds of the sample (71%) could describe three government-recommended prevention methods. Differences in knowledge of symptoms and preventative measures by sociodemographic factors are shown in Table 2. After controlling for other sociodemographic factors, participants with inadequate health literacy were significantly less likely (p<0.001) to be able to name three key symptoms of COVID-19, and less likely to be able to report three preventative methods (p<0.001).

Participants reported spending on average 1.3 hours (SD 1.3) per day getting news or learning about COVID-19. The three most frequently endorsed sources of information were public television (67.6%), social media (64.4%) and government websites (63.9%). Sociodemographic disparities in understanding government messaging was evident (Table 2); younger participants, females, those with inadequate health literacy, poorer general health, primarily speaking English at home, and those without private health insurance reporting greater difficulty understanding. After adjusting for other sociodemographic factors, individuals with inadequate health literacy reported significantly more difficulty (p<0.001) understanding government messaging in relation to COVID-19.

### Behaviour change and impact

Changes to plans due to COVID-19 were reported by the vast majority (92%) of the sample, with 84% agreeing that COVID-19 has impacted their daily routine. Only 9% of the total sample felt “not at all” personally prepared for widespread outbreak of COVID-19. Compared to before the introduction of COVID-19 restrictions, 26% reported drinking more alcohol, and 37% reported eating more unhealthy snacks. Variations in behaviour change and impact as a function of sociodemographic factors are shown in Table 2.

After controlling for other sociodemographic factors (Table 3), individuals with inadequate health literacy were less likely (p = 0.014) to have made changes to their plans, and less likely to report social distancing as important (p<0.001), but much more likely to feel personally unprepared for widespread outbreak (p<0.001) compared to those with adequate health literacy. Individuals with inadequate health literacy were less likely to have increased their alcohol intake (p = 0.013), but not eating unhealthy snacks (p = 0.20) compared to those with adequate health literacy. Of those taking regular prescription medication, individuals with inadequate health literacy reported it was more difficult to remember and access medication during lockdown (p = 0.013).

### Support for misinformation

Across the sample, support for misinformation about COVID-19 was generally more prominent in younger age groups, males, those with inadequate health literacy, fewer chronic health conditions, and those who spoke a LOTE at home (Table 2). After controlling for other sociodemographic factors (Table 3), those with inadequate health literacy were significantly more likely to agree that the effectiveness of vaccines is made up (p<0.001), the threat of COVID-19 is greatly exaggerated (p = 0.029), the benefits of herd immunity for COVID-19 are being covered up (p = 0.022), and that government restrictions are stronger than what is needed (p = 0.003), compared to individuals with adequate health literacy.

## DISCUSSION

Our findings show important disparities in knowledge, attitudes, beliefs and behaviours related to COVID-19 that have the potential to undermine efforts to reduce viral transmission and may lead to social inequalities in health outcomes in Australia. People with lower health literacy had poorer understanding of COVID-19 symptoms, were less able to identify behaviours to prevent infection, and experienced more difficulty finding information and understanding government messaging about COVID-19. They were less likely to rate social distancing as important and reported more difficulty remembering and accessing medication since lockdown. They felt less prepared and more anxious about COVID-19, reported experiencing greater financial toxicity but also perceived they were less likely to get sick from COVID-19. Similar patterns were observed among those who primarily spoke a LOTE at home. Notably, there was markedly higher endorsement of misinformation beliefs in people with lower health literacy and LOTE background, which is a concern as these items relate to ongoing efforts to prevent viral transmission and trust in vaccination –the major hope for mitigating COVID-19 worldwide.

The findings support our earlier concerns about the low level of attention paid to health literacy in COVID-19 public health messaging (6). In our preliminary analysis of health information presented on government websites we found readability scores to be higher than the level suitable for the average Australian (reading Grade 8), and far higher than the grade required for low literacy communities (Grade 5) including those with English as a second language. The findings echo results reported in the US (Chicago) sister survey conducted in March (7). Here, even larger social disparities in key knowledge, attitude and prevention behaviours were reported. Although Australia is in a much more favourable position in relation to COVID-19 than the US for many reasons, the need for ongoing attention to social variation in community uptake of public health messages remains. Until an effective vaccine is available, our primary defence against the spread of COVID-19 is behaviour change. Effective behaviour change relies on diverse communities and patient groups being able to understand, trust and act on evolving health advice. Our comprehensive survey of over 4000 Australians suggests there are important knowledge and attitude gaps which may threaten efforts to reduce viral transmission in Australia.

A systematic review of communication during the H1N1 (swine flu) pandemic in 118 studies (17) reported a consistent association between social inequalities in communication and emergency preparedness outcomes. Trust in sources of information, worry and levels of knowledge about the disease, and routine media exposure as well as information-seeking behaviours, were related to greater likelihood of adoption of recommended viral infection prevention behaviours. The review suggests that when inequalities are addressed in communication the effectiveness of the pandemic response can be increased.

## Limitations of study

The recruited sample is large and diverse but is not statistically representative of the Australian population. The proportion of Australians from non-English speaking backgrounds was small and we had very few Aboriginal and Torres Strait Islander participants in our sample (2%), although this similar to the national estimate of 3.5%. Future surveys need to target these groups as we were unable to do so with limited time and funds for this project. Our sample of adults with low health literacy is similar to other studies we have conducted (∼15%)(18). The single item measure of health literacy used for assessment is simple and non-stigmatising to administer, however it under-reports problems with health literacy, identifying only those with very low health literacy levels (19). It was correlated with our additional related measure of numeracy, ehealth literacy and graphical literacy (all P values < 0.001). Future studies should use more comprehensive literacy measures to understand the associations with key knowledge, attitude and behaviour outcomes related to COVID-19.

## Conclusion

COVID-19 presents a disproportionate burden to people with chronic disease, who are also more likely to have poorer health literacy and speak a language other than English at home. Health messages must be tailored to meet the needs of these groups as our study shows important disparities in understanding, beliefs and behaviours that may put already vulnerable people at greater risk. Those with the greatest burden of chronic disease are most disadvantaged, and most likely to experience severe disease and die from COVID-19. It is imperative that health advice reaches them in a way they understand and can implement. Addressing the health literacy needs of the community in public health messaging about COVID-19 must now be a priority in Australia.

## Data Availability

The data may be available upon request from the authors.

## Box 1.

**The known**

- People with chronic disease are more susceptible to severe illness and death from COVID-19; the same groups frequently have lower health literacy.

**The new**

- First national data on variations in knowledge, attitudes and uptake of public health messages by health literacy in Australia.
- It shows important disparities with poorer outcomes by health literacy and primary language spoken at home.

**The implications**

- Inadequate understanding and uptake of behavioural advice may undermine public health efforts to reduce viral transmission.
- Health messages must be tailored to meet the needs of diverse populations or may put already vulnerable people at greater risk.

## Acknowledgements

We would like to acknowledge the members of the Australian public who participated in this survey and Hilary Cox for her administrative assistance in the preparation of this article.

No funder had any role in the study.

There are no conflicts of interest.

All authors had full access to all of the data including statistical reports and tables.

A copy of all measures in included in the Appendix.

---

**Supplementary Table S1.**
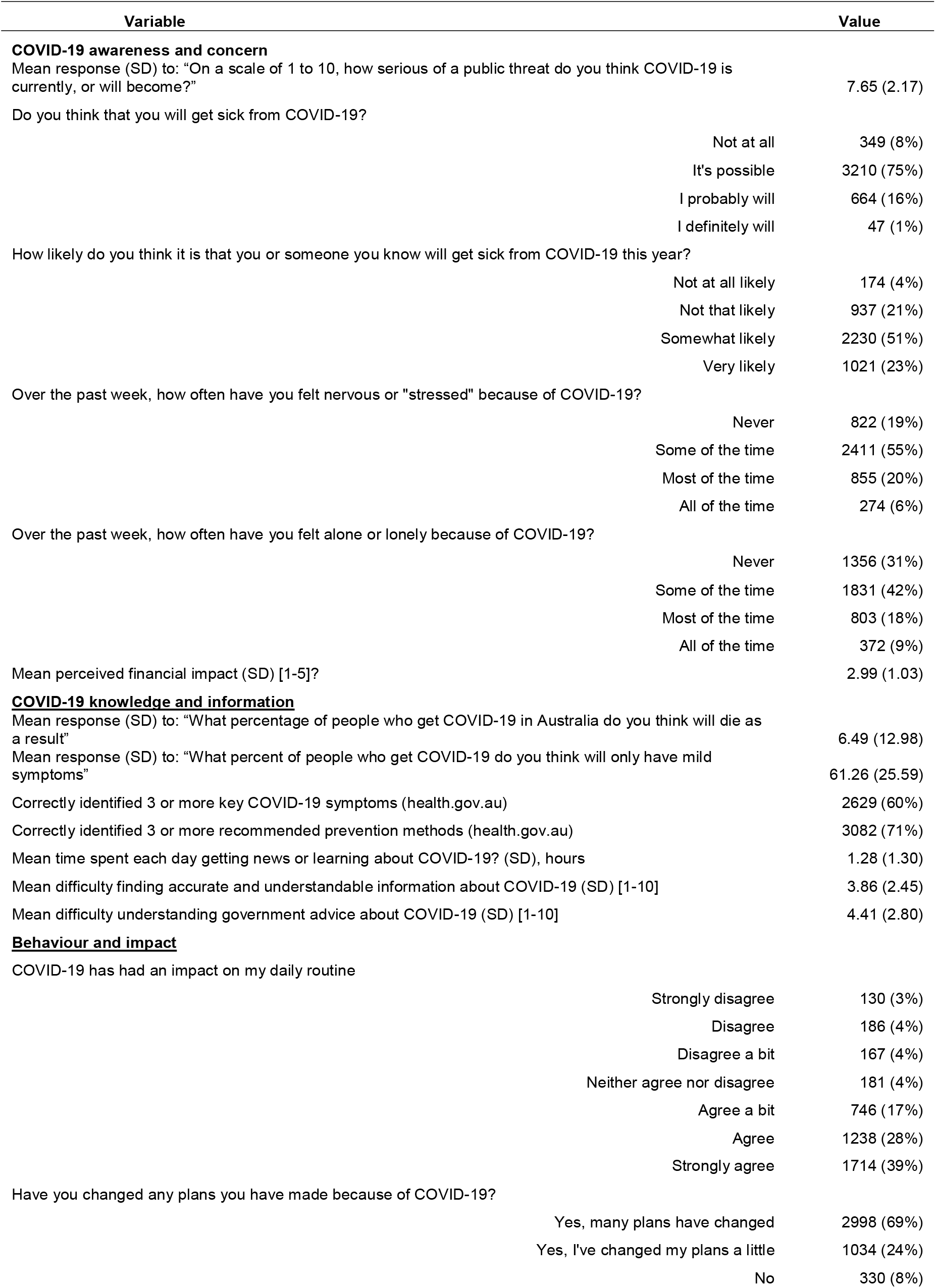
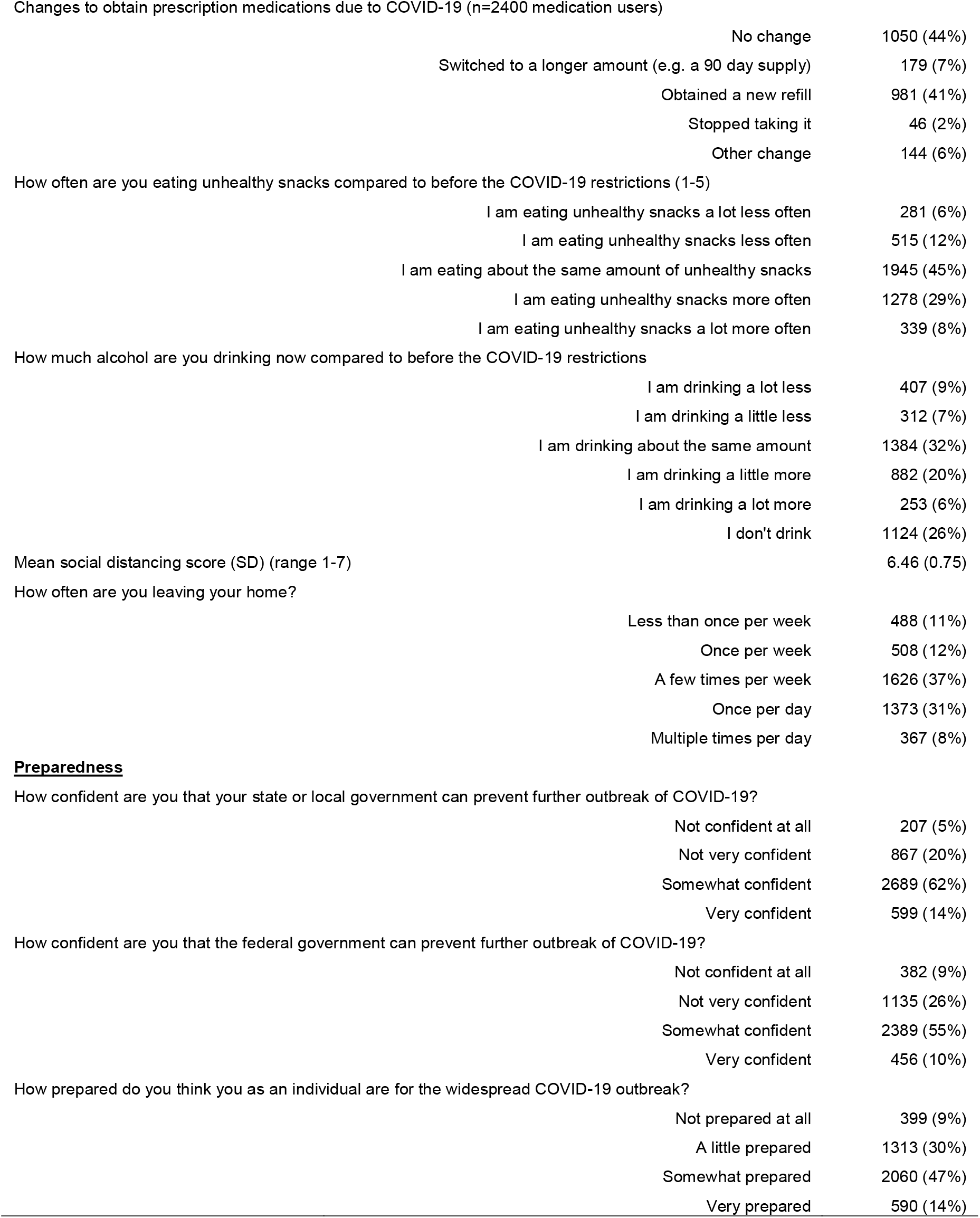
Knowledge, attitudes, beliefs and behaviours related to COVID-19 in full analysis sample (N=4362).

| Variable | Value |
| --- | --- |
| **COVID-19 awareness and concern** |  |
| Mean response (SD) to: "On a scale of 1 to 10, how serious of a public threat do you think COVID-19 is currently, or will become?" | 7.65 (2.17) |
| Do you think that you will get sick from COVID-19? |  |
| Not at all | 349 (8%) |
| It's possible | 3210 (75%) |
| I probably will | 664 (16%) |
| I definitely will | 47 (1%) |
| How likely do you think it is that you or someone you know will get sick from COVID-19 this year? |  |
| Not at all likely | 174 (4%) |
| Not that likely | 937 (21%) |
| Somewhat likely | 2230 (51%) |
| Very likely | 1021 (23%) |
| Over the past week, how often have you felt nervous or "stressed" because of COVID-19? |  |
| Never | 822 (19%) |
| Some of the time | 2411 (55%) |
| Most of the time | 855 (20%) |
| All of the time | 274 (6%) |
| Over the past week, how often have you felt alone or lonely because of COVID-19? |  |
| Never | 1356 (31%) |
| Some of the time | 1831 (42%) |
| Most of the time | 803 (18%) |
| All of the time | 372 (9%) |
| Mean perceived financial impact (SD) [1-5]? | 2.99 (1.03) |
| **COVID-19 knowledge and information** |  |
| Mean response (SD) to: "What percentage of people who get COVID-19 in Australia do you think will die as a result" | 6.49 (12.98) |
| Mean response (SD) to: "What percent of people who get COVID-19 do you think will only have mild symptoms" | 61.26 (25.59) |
| Correctly identified 3 or more key COVID-19 symptoms (health.gov.au) | 2629 (60%) |
| Correctly identified 3 or more recommended prevention methods (health.gov.au) | 3082 (71%) |
| Mean time spent each day getting news or learning about COVID-19? (SD), hours | 1.28 (1.30) |
| Mean difficulty finding accurate and understandable information about COVID-19 (SD) [1-10] | 3.86 (2.45) |
| Mean difficulty understanding government advice about COVID-19 (SD) [1-10] | 4.41 (2.80) |
| **Behaviour and impact** |  |
| COVID-19 has had an impact on my daily routine |  |
| Strongly disagree | 130 (3%) |
| Disagree | 186 (4%) |
| Disagree a bit | 167 (4%) |
| Neither agree nor disagree | 181 (4%) |
| Agree a bit | 746 (17%) |
| Agree | 1238 (28%) |
| Strongly agree | 1714 (39%) |
| Have you changed any plans you have made because of COVID-19? |  |
| Yes, many plans have changed | 2998 (69%) |
| Yes, I've changed my plans a little | 1034 (24%) |
| No | 330 (8%) |

|  |  |  |
| --- | --- | --- |
| Changes to obtain prescription medications due to COVID-19 (n=2400 medication users) |  |  |
|  | No change | 1050 (44%) |
|  | Switched to a longer amount (e.g. a 90 day supply) | 179 (7%) |
|  | Obtained a new refill | 981 (41%) |
|  | Stopped taking it | 46 (2%) |
|  | Other change | 144 (6%) |
| How often are you eating unhealthy snacks compared to before the COVID-19 restrictions (1-5) |  |  |
|  | I am eating unhealthy snacks a lot less often | 281 (6%) |
|  | I am eating unhealthy snacks less often | 515 (12%) |
|  | I am eating about the same amount of unhealthy snacks | 1945 (45%) |
|  | I am eating unhealthy snacks more often | 1278 (29%) |
|  | I am eating unhealthy snacks a lot more often | 339 (8%) |
| How much alcohol are you drinking now compared to before the COVID-19 restrictions |  |  |
|  | I am drinking a lot less | 407 (9%) |
|  | I am drinking a little less | 312 (7%) |
|  | I am drinking about the same amount | 1384 (32%) |
|  | I am drinking a little more | 882 (20%) |
|  | I am drinking a lot more | 253 (6%) |
|  | I don't drink | 1124 (26%) |
| Mean social distancing score (SD) (range 1-7) |  | 6.46 (0.75) |
| How often are you leaving your home? |  |  |
|  | Less than once per week | 488 (11%) |
|  | Once per week | 508 (12%) |
|  | A few times per week | 1626 (37%) |
|  | Once per day | 1373 (31%) |
|  | Multiple times per day | 367 (8%) |
| **<u>Preparedness</u>** |  |  |
| How confident are you that your state or local government can prevent further outbreak of COVID-19? |  |  |
|  | Not confident at all | 207 (5%) |
|  | Not very confident | 867 (20%) |
|  | Somewhat confident | 2689 (62%) |
|  | Very confident | 599 (14%) |
| How confident are you that the federal government can prevent further outbreak of COVID-19? |  |  |
|  | Not confident at all | 382 (9%) |
|  | Not very confident | 1135 (26%) |
|  | Somewhat confident | 2389 (55%) |
|  | Very confident | 456 (10%) |
| How prepared do you think you as an individual are for the widespread COVID-19 outbreak? |  |  |
|  | Not prepared at all | 399 (9%) |
|  | A little prepared | 1313 (30%) |
|  | Somewhat prepared | 2060 (47%) |
|  | Very prepared | 590 (14%) |

**Supplementary Table S2.** Multivariable regression models exploring variation in awareness and concern outcomes by sociodemographic factors. Continuous outcomes are presented as estimated marginal means (95% confidence intervals). Values in bold type have statistical evidence (p< 0.05) of a main effect (see key for p-value level). Categorical outcomes are presented as adjusted relative risks (95% confidence intervals). Level of statistical evidence for main effects are indicated by the symbol provided accompanying the reference value; significant contrasts relative to the reference group are indicated in bold type.

|  |  | Awareness and concern |  |  |  |  |
| --- | --- | --- | --- | --- | --- | --- |
|  |  | Mean Seriousness of threat<br>(95% CI) | Not nervous/<br>Stressed<br>Risk Ratio (95% CI) | Not likely to get sick<br>Risk Ratio (95% CI) | Mean STAI<br>(95% CI) | Mean Financial Toxicity<br>(95% CI) |
| **Age group** |  |  |  |  |  |  |
| | 18 to 25 years | **7.41 (7.27-7.56)\$** | 1.00 (ref)\$ | 1.00 (ref)\$ | **48.15 (47.20-49.10)\$** | **3.19 (3.13-3.26)\$** |
|  | 26 to 40 years | **7.47 (7.34-7.59)** | 0.83 (0.67-1.03) | 1.12 (0.96-1.32) | **47.21 (46.38-48.04)** | **3.17 (3.12-3.23)** |
|  | 41 to 55 years | **7.74 (7.60-7.88)** | 1.19 (0.96-1.47) | 1.16 (0.97-1.38) | **43.10 (42.17-44.02)** | **3.03 (2.96-3.09)** |
| | 56 to 90 years | **7.92 (7.79-8.05)** | **1.95 (1.63-2.33)\$** | **1.73 (1.48-2.01)\$** | **36.25 (35.37-37.12)** | **2.60 (2.54-2.66)** |
| **Gender** |  |  |  |  |  |  |
| | Male | **7.31 (7.21-7.42)\$** | 1.00 (ref) | 1.00 (ref) | **41.34 (40.65-42.03)\$** | 2.98 (2.93-3.03) |
| | Female | **7.85 (7.77-7.94)** | **0.52 (0.46-0.58)\$** | **0.71 (0.64-0.78)\$** | **44.83 (44.27-45.38)** | 2.99 (2.95-3.03) |
| **Health Literacy** |  |  |  |  |  |  |
| | Inadequate | **7.28 (7.09-7.46)\$** | 0.85 (0.70-1.03) | **1.19 (1.03-1.37)^** | **47.11 (45.57-48.36)\$** | **3.18 (3.10-3.27)\$** |
|  | Adequate | **7.69 (7.63-7.76)** | 1.00 (ref) | 1.00 (ref) | **42.93 (42.45-43.39)** | **2.96 (2.92-2.99)** |
| **Chronic Health Conditions** |  |  |  |  |  |  |
| | None | **7.53 (7.44-7.62)\$** | 1.00 (ref)\$ | 1.00 (ref)\$ | **42.60 (41.99-43.21)\$** | **2.95 (2.91-3.00)^** |
|  | One | **7.62 (7.50-7.74)** | 0.91 (0.79-1.05) | **0.88 (0.78-0.99)^** | **43.47 (42.66-44.27)** | **2.98 (2.92-3.03)** |
| | Two or more | **7.96 (7.82-8.11)** | **0.71 (0.60-0.84)\$** | **0.73 (0.63-0.85)\$** | **45.64 (44.65-46.64)** | **3.07 (3.01-3.14)** |
| **Language at home** |  |  |  |  |  |  |
| | English | 7.64 (7.57-7.70) | 1.00 (ref) | 1.00 (ref) | 43.48 (43.04-43.93) | **2.97 (2.94-3.00)\$** |
| | Other | 7.69 (7.43-7.95) | **0.72 (0.53-0.99)^** | **1.46 (1.22-1.74)\$** | 43.02 (41.28-44.76) | **3.24 (3.12-3.36)** |
| **Private health insurance** |  |  |  |  |  |  |
|  | Yes | **7.70 (7.63-7.79)#** | 1.00 (ref) | 1.00 (ref) | 43.17 (42.63-43.71) | **2.95 (2.91-2.99)#** |
| | No/Unknown | **7.53 (7.42-7.63)** | **1.18 (1.04-1.33)#** | **1.21 (1.10-1.35)\$** | 43.95 (43.24-44.67) | **3.05 (3.00-3.09)** |
| **Employment** |  |  |  |  |  |  |
| | Paid employment | 7.64 (7.54-7.74) | 1.00 (ref) | 1.00 (ref) | **42.68 (42.07-43.30)\$** | **2.82 (2.78-2.87)\$** |
| | Other | 7.64 (7.55-7.74) | **1.20 (1.05-1.37)#** | **1.24 (1.10-1.38)\$** | **44.30 (43.65-44.94)** | **3.16 (3.12-3.20)** |

**^P<0.05; #P<0.01; \$P<0.001**

**Supplementary Table S3.**
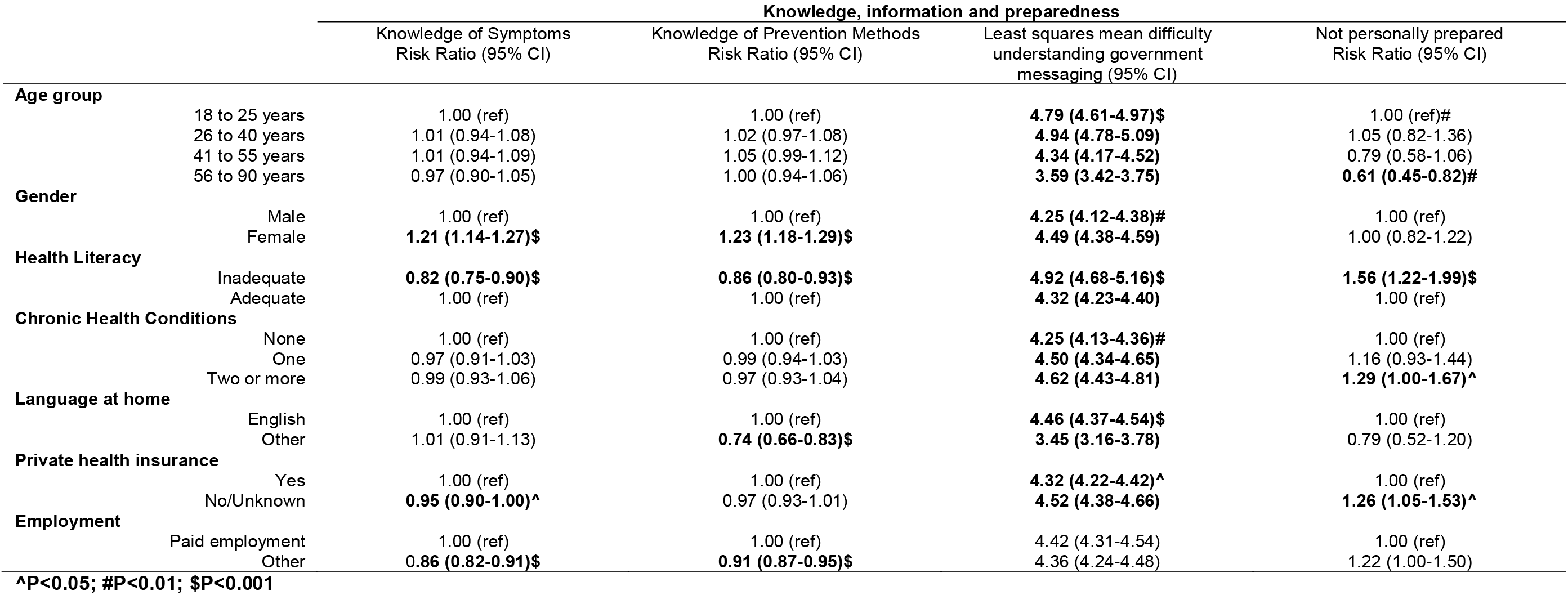
Multivariable regression models exploring variation in knowledge, information and preparedness by sociodemographic factors. Continuous outcomes are presented as estimated marginal means (95% confidence intervals). Values in bold type have statistical evidence (p< 0.05) of a main effect (see key for p-value level). Categorical outcomes are presented as adjusted relative risks (95% confidence intervals). Level of statistical evidence for main effects are indicated by the symbol provided accompanying the reference value; significant contrasts relative to the reference group are indicated in bold type.

|  |  | Knowledge, information and preparedness |  |  |  |
| --- | --- | --- | --- | --- | --- |
|  |  | Knowledge of Symptoms<br>Risk Ratio (95% CI) | Knowledge of Prevention Methods<br>Risk Ratio (95% CI) | Least squares mean difficulty<br>understanding government<br>messaging (95% CI) | Not personally prepared<br>Risk Ratio (95% CI) |
| **Age group** |  |  |  |  |  |
| | 18 to 25 years | 1.00 (ref) | 1.00 (ref) | **4.79 (4.61-4.97)\$** | 1.00 (ref)# |
|  | 26 to 40 years | 1.01 (0.94-1.08) | 1.02 (0.97-1.08) | **4.94 (4.78-5.09)** | 1.05 (0.82-1.36) |
|  | 41 to 55 years | 1.01 (0.94-1.09) | 1.05 (0.99-1.12) | **4.34 (4.17-4.52)** | 0.79 (0.58-1.06) |
|  | 56 to 90 years | 0.97 (0.90-1.05) | 1.00 (0.94-1.06) | **3.59 (3.42-3.75)** | **0.61 (0.45-0.82)#** |
| **Gender** |  |  |  |  |  |
|  | Male | 1.00 (ref) | 1.00 (ref) | **4.25 (4.12-4.38)#** | 1.00 (ref) |
| | Female | **1.21 (1.14-1.27)\$** | **1.23 (1.18-1.29)\$** | **4.49 (4.38-4.59)** | 1.00 (0.82-1.22) |
| **Health Literacy** |  |  |  |  |  |
| | Inadequate | **0.82 (0.75-0.90)\$** | **0.86 (0.80-0.93)\$** | **4.92 (4.68-5.16)\$** | **1.56 (1.22-1.99)\$** |
|  | Adequate | 1.00 (ref) | 1.00 (ref) | **4.32 (4.23-4.40)** | 1.00 (ref) |
| **Chronic Health Conditions** |  |  |  |  |  |
|  | None | 1.00 (ref) | 1.00 (ref) | **4.25 (4.13-4.36)#** | 1.00 (ref) |
|  | One | 0.97 (0.91-1.03) | 0.99 (0.94-1.03) | **4.50 (4.34-4.65)** | 1.16 (0.93-1.44) |
|  | Two or more | 0.99 (0.93-1.06) | 0.97 (0.93-1.04) | **4.62 (4.43-4.81)** | **1.29 (1.00-1.67)^** |
| **Language at home** |  |  |  |  |  |
| | English | 1.00 (ref) | 1.00 (ref) | **4.46 (4.37-4.54)\$** | 1.00 (ref) |
| | Other | 1.01 (0.91-1.13) | **0.74 (0.66-0.83)\$** | **3.45 (3.16-3.78)** | 0.79 (0.52-1.20) |
| **Private health insurance** |  |  |  |  |  |
|  | Yes | 1.00 (ref) | 1.00 (ref) | **4.32 (4.22-4.42)^** | 1.00 (ref) |
|  | No/Unknown | **0.95 (0.90-1.00)^** | 0.97 (0.93-1.01) | **4.52 (4.38-4.66)** | **1.26 (1.05-1.53)^** |
| **Employment** |  |  |  |  |  |
|  | Paid employment | 1.00 (ref) | 1.00 (ref) | 4.42 (4.31-4.54) | 1.00 (ref) |
| | Other | **0.86 (0.82-0.91)\$** | **0.91 (0.87-0.95)\$** | 4.36 (4.24-4.48) | 1.22 (1.00-1.50) |

**^P<0.05; #P<0.01; \$P<0.001**

**Supplementary Table S4.**
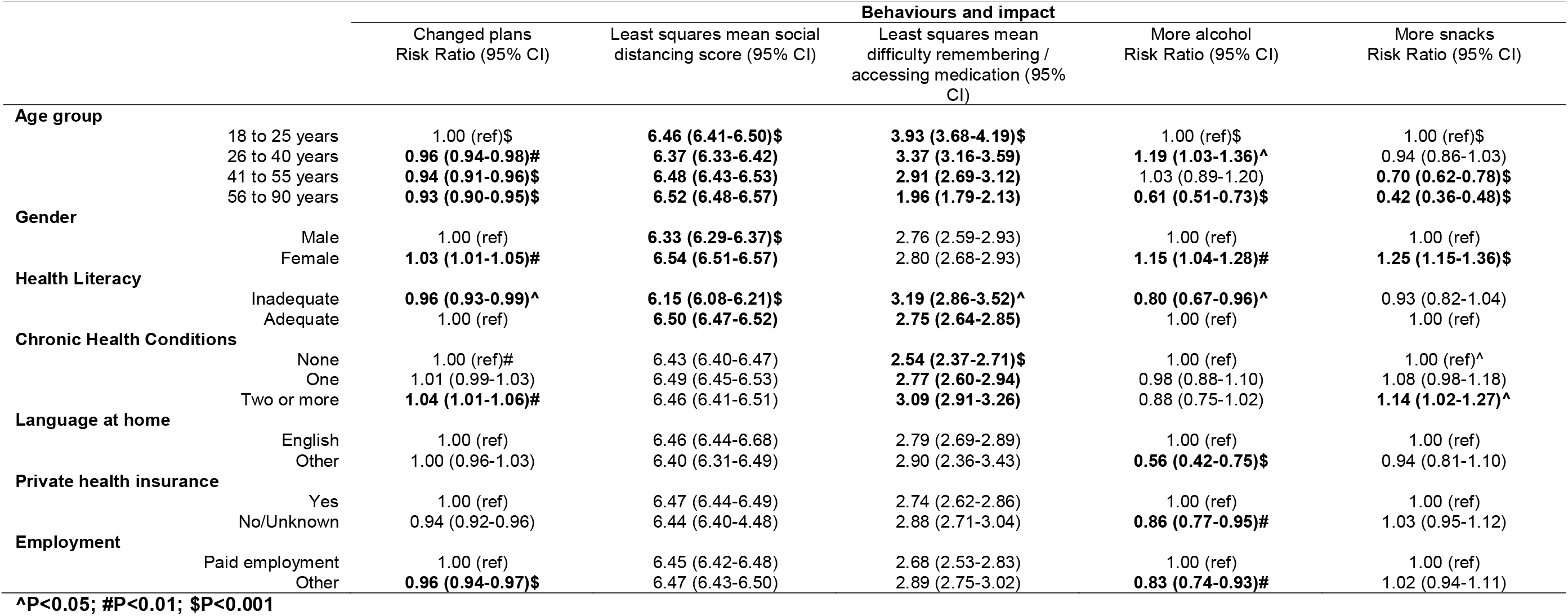
Multivariable regression models exploring variation in behaviour change and impact by sociodemographic factors. Continuous outcomes are presented as estimated marginal means (95% confidence intervals). Values in bold type have statistical evidence (p< 0.05) of a main effect (see key for p-value level). Categorical outcomes are presented as adjusted relative risks (95% confidence intervals). Level of statistical evidence for main effects are indicated by the symbol provided accompanying the reference value; significant contrasts relative to the reference group are indicated in bold type.

**Supplementary Table S5.** Multivariable regression models exploring variation in pseudoscience and conspiracy beliefs by sociodemographic factors. Continuous outcomes are presented as estimated marginal means (95% confidence intervals). Values in bold type have statistical evidence (p< 0.05) of a main effect (see key for p-value level). Categorical outcomes are presented as adjusted relative risks (95% confidence intervals). Level of statistical evidence for main effects are indicated by the symbol provided accompanying the reference value; significant contrasts relative to the reference group are indicated in bold type.

|  |  | COVID-19/ Vaccination Misinformation Beliefs |  |  |  |
| --- | --- | --- | --- | --- | --- |
|  |  | Vaccine effectiveness made-up<br>Risk Ratio (95% CI) | COVID-19 threat greatly<br>exaggerated<br>Risk Ratio (95% CI) | Herd immunity beneficial and is<br>covered up<br>Risk Ratio (95% CI) | Government restrictions stronger<br>than needed<br>Risk Ratio (95% CI) |
| **Age group** | 18 to 25 years | 1.00 (ref) | 1.00 (ref)\$ | 1.00 (ref)\$ | 1.00 (ref)# |
|  | 26 to 40 years | 1.06 (0.88-1.27) | 0.90 (0.75-1.08) | **0.77 (0.64-0.92)#** | **1.43 (1.16-1.77)#** |
| | 41 to 55 years | 1.01 (0.82-1.24) | **0.62 (0.49-0.78)\$** | **0.62 (0.50-0.78)\$** | 1.17 (0.92-1.49) |
| | 56 to 90 years | 0.92 (0.75-1.12) | **0.37 (0.28-0.78)\$** | **0.46 (0.36-0.57)\$** | 0.99 (0.78-1.28) |
| **Gender** | Male | 1.00 (ref) | 1.00 (ref) | 1.00 (ref) | 1.00 (ref) |
| | Female | **0.85 (0.74-0.97)^** | **0.55 (0.48-0.64)\$** | **0.74 (0.64-0.85)\$** | **0.61 (0.53-0.71)\$** |
| **Health Literacy** | Inadequate | **1.45 (1.22-1.72)\$** | **1.24 (1.02-1.50)^** | **1.25 (1.03-1.51)^** | **1.35 (1.11-1.65)#** |
|  | Adequate | 1.00 (ref) | 1.00 (ref) | 1.00 (ref) | 1.00 (ref) |
| **Chronic Health Conditions** | None | 1.00 (ref) | 1.00 (ref) | 1.00 (ref) | 1.00 (ref)^ |
|  | One | 1.09 (0.93-1.27) | 0.89 (0.74-1.06) | 1.00 (0.85-1.18) | **0.80 (0.66-0.96)^** |
|  | Two or more | 1.07 (0.89-1.28) | 0.88 (0.70-1.11) | 0.89 (0.71-1.10) | 0.84 (0.68-1.05) |
| **Language at home** | English | 1.00 (ref) | 1.00 (ref) | 1.00 (ref) | 1.00 (ref) |
| | Other | **1.79 (1.48-2.18)\$** | **1.33 (1.05-1.67)^** | 1.04 (0.80-1.35) | 1.21 (0.93-1.58) |
| **Private health insurance** | Yes | 1.00 (ref) | 1.00 (ref) | 1.00 (ref) | 1.00 (ref) |
|  | No/Unknown | **1.18 (1.03-1.35)^** | 1.09 (0.93-1.27) | 0.89 (0.76-1.03) | 1.06 (0.91-1.24) |
| **Employment** | Paid employment | 1.00 (ref) | 1.00 (ref) | 1.00 (ref) | 1.00 (ref) |
|  | Other | **1.16 (1.00-1.33)^** | 1.02 (0.87-1.20) | 1.05 (0.90-1.22) | 1.00 (0.84-1.18) |

^ $P < 0.05$ ; # $P < 0.01$ ; \$ $P < 0.001$

## Notes

### Competing Interest Statement

The authors have declared no competing interest.

### Funding Statement

No external funding received.

### Author Declarations

University of Sydney Human Ethics Committee Project number: 2020/212

